# Gender and Socio-Economic Status as moderators in the associations between Social Support Sources and Adolescents’ Mental and Behavioral Health Indicators

**DOI:** 10.1101/2023.08.31.23294835

**Authors:** E. Bermejo-Martins, M. Torres-Sahli, K. Rich Madsen, M. Tabs-Damgraads, L. Nielsen, C.B. Meilstrup, M. Toftager, Z. I. Santini

## Abstract

**Background:** Extensive research has established the intricate links between diverse social support sources and vital adolescent health indicators, such as mental wellbeing (MWB), problematic social media use (PSMU), and physical activity (PA). However, existing studies have not explored these interrelationships within a unified model or examined the moderating effects of gender and socio-economic status (SES).

**Methods:** This cross-sectional study employed a representative Danish sample of 2.034 adolescents, aged 13 and 15 years. A Multi-group Structural Equation Model (SEM) and covariance-based comparisons analysis utilized items from Multidimensional Scales of Perceived Social Support, The Warwick–Edinburgh Mental Well-being Scale, Social Media Disorder Scale, and measures of PA duration and frequency.

**Results:** The associations between MWB-Teacher Support and MWB-Classmate Support were stronger in low SES adolescents than those with mid-high SES. Notably, the PSMU-Family Support negative association was more pronounced among girls, while PSMU-Friend Support’s negative relationship was stronger among boys. The PA-Family Support positive relationship was more robust in boys, while the PA-Teacher Support positive association was stronger among low-mid SES adolescents than those in high SES. The positive correlation between PA and MWB was stronger among boys and mid-low SES adolescents.

**Conclusions:** Strategies designed to enhance family and school support, considering gender and SES, could effectively promote MWB and deter behavioural issues like PSMU and sedentary behaviours in adolescents.

## 1. Introduction

The mental health of today’s youth is increasingly precarious, a growing public health concern (Fusar-Poli et al., 2021). A marked decline in adolescent mental health, particularly among girls, is evident over the past decade (Bor et al., 2014; Potrebny, Wilium & Lundegård, 2017). The Covid-19 pandemic further exacerbated mental health issues worldwide, including anxiety, depression, and suicidal ideation (Oliveira et al., 2022; Samji et al., 2022).

The salutogenic view posits mental health as more than mere absence of mental health issues. Emphasizing positive mental health aspects extends beyond a purely risk reduction approach. Aspects such as purpose in life, sound social relations, and a sense of contribution significantly affect mental health (Koushede & Donoan, 2022).

Critical indicators of adolescent mental and behavioral health include perceived social support, PSMU, PA, and MWB (Khan et al., 2021; Jakobsen et al., 2022; Maenhout et al., 2020). Different social support sources significantly influence adolescent mental health (Chu, Saucier & Hafner, 2010; Herrde & Hemphill, 2018; Brooks et al., 2015), with social support associated with positive mental health indicators like hope, meaningfulness, and subjective well-being (Jakobsen et al., 2022). In contrast, inadequate social support may detrimentally impact mental well-being and increase the likelihood of PSMU (Walsh et al., 2020) and sedentary behaviors (Jusiené et al., 2022; Hu et al., 2021).

An increasingly recognized issue is the potentially damaging influence of social media on adolescent mental health (Twenge et al., 2018). This issue is partly due to social isolation and self-esteem issues among adolescents exhibiting PSMU (Ivie et al., 2020; Kelly et al., 2018; Raudseppn & Kais, 2019; Scott & Woods, 2018).

Previous research has identified gender and SES as determinants of adolescent perceived social support (Van Droogenbroeck, Spruyt & Keppens, 2018; Gomes et al., 2020), MWB (Nagy-Pénzes; Vincze & Bíró, 2020; Maheswari, K., & Martin,2022; Yoon et al., 2022), PSMU (Raudsepp et al., 2019; Boniel-Nissim et al., 2022; Micheli, 2016), and PA (Hu et al, 2021; Humbert et al., 2006).

However, an analysis of the moderating effect of gender and SES on these variables’ interrelationships is lacking. It’s vital to discern how varying support sources may differentially affect adolescent health outcomes.

This study seeks to elucidate the relationships between social support sources, MWB, PSMU, and PA, while accounting for the role of gender and SES. We posit that social support sources will associate positively with MWB and PA, and negatively with PSMU. PSMU levels will negatively relate to MWB and PA, and PA will positively link with MWB. Furthermore, we expect that the strength of these associations varies depending on gender and SES groups. Given the paucity of literature, we do not propose a specific moderation effect.

## 2. Methods

### Participants and design

The present study uses data from the Danish contribution to the international study titled “Health Behaviour in School-aged Children” (HBSC). The HBSC is a school-based investigation into adolescent health, their health behaviors, and social determinants. It samples school-aged adolescents in the 5th, 7th, and 9th grades (equivalent to ages 11, 13, and 15, respectively) every four years, following an international standardized methodology which involves standard procedures for sampling and item translation (Inchley et al., 2020).

Our study adopted a cross-sectional design, using data from the 2018 Danish HBSC survey, which included only 7th and 9th graders, or 13-and 15-year-olds. Our sample comprised 2.034 Danish adolescents, with an even gender distribution of males (n=1032, 50.7%) and females (n=1002, 49.3%) (refer to Tables 1 and 2).

**Table 1.** Characteristics of the sample according to main variables and group.

| *Variable* | *Gender* |  | *p value*<br>† | *SES* |  |  | *p value*<br>¥ |
| --- | --- | --- | --- | --- | --- | --- | --- |
|  | **Male** | **Female** |  | **High** | **Middle** | **Low** |  |
| ***Family Support (n)*** | 1010 | 990 |  | 790 | 799 | 411 |  |
| *Median (IQR)* | 19 (16-20) | 18 (16-20) | .012 | 19 (16-20) | 18 (16-20) | 18 (16-20) | .003 |
| *missing* | 22 (2%) | 12 (1%) |  | 12 (1%) | 10 (1%) |  |  |
| ***Friends Support (n)*** | 1004 | 991 |  | 790 | 795 | 410 |  |
| *Median (IQR)* | 25 (22-28) | 26 (22-28) | .016 | 26 (23-28) | 26 (22-28) | 25 (21-28) | .097 |
| *missing* | 28 (3%) | 11(1%) |  | 12 (1%) | 14 (2%) | 13 (3%) |  |
| ***Classmates Support (n)*** | 1022 | 997 |  | 796 | 803 | 420 |  |
| *Median (IQR)* | 12 (11-14) | 12 (10-13) | <.001 | 12 (11-14) | 12 (11-13) | 12 (10-14) | .007 |
| *missing* | 10 (1%) | 5 (0.5%) |  | 6 (1%) | 6 (1%) | 3 (1%) |  |
| ***Teacher Support (n)*** | 1022 | 1001 |  | 797 | 806 | 420 |  |
| *Median (IQR)* | 12 (11-14) | 12(12-13) | < .001 | 12 (10-14) | 12 (10-13) | 12 (10-14) | .006 |
| *missing* | 10 (1%) | 1 (0.1%) |  | 5 (1%) | 3 (0.4%) | 3 (1%) |  |
| ***Mental Well-being (n)*** | 923 | 942 |  | 742 | 756 | 367 |  |
| *Median (IQR)* | 28 (27-31) | 28 (25-29) | < .001 | 28 (26-30) | 28 (26-30) | 28 (26-30) | .275 |
| *missing* | 109 (11%) | 60 (6%) |  | 60 (7%) | 53 (7%) | 56 (13%) |  |
| ***PSMU (n)*** | 968 | 974 |  | 774 | 782 | 386 |  |
| *Median (IQR)* | 9 (9-11) | 10 (9-12) | < .001 | 10 (9-11) | 10 (9-11) | 10 (9-12) | .032 |
| *missing* | 64 (6%) | 28 (3%) |  | 28 (3%) | 27 (3%) | 37 (9%) |  |
| ***Physical Activity (n)*** | 956 | 960 |  | 756 | 778 | 382 |  |
| *Median (IQR)* | 17 (14-20) | 16 (13-19) | < 0.001 | 17 (14-20) | 16 (13-20) | 16 (12-19) | <0.001 |
| *missing* | 76 (7%) | 42 (4%) |  | 46 (6%) | 31(4%) | 41 (10%) |  |

PSMU= Problematic Social Media Use; †=Mann-Whitney test, ¥=The Kruskal–Wallis’s test

**Table 2.** Models’ fit and measurement invariance by Gender and SES groups.

| Model | Observation<br>(missing<br>patterns) | Model fit | $\chi^2$ (Df) | RMSEA | SRMR | CFI | Invariance |
| --- | --- | --- | --- | --- | --- | --- | --- |
| **1.Gender**<br>**(all)** | 2034 (87) | Good | 1998.8(506) | .038 | .050 | .98 |  |
| *Female* | 1002 (43) | Acceptable | 1330.3(506) | .040 | .061 | .98 |  |
| *Male* | 1032 (69) | Good | 1085.7(506) | .033 | .052 | .98 |  |
| **1.Configural invariance** |  | Good | 2389.3 (1012) | .034 | .042 | .96 | Yes |
| **2.Metric invariance** |  | Good | 2389.3 (1012) | .034 | .042 | .96 | Yes |
| **3.Scalar invariance** |  | Poor | 2479.6 (1039) ** | .034 | .044 | .95 | No |
| **4.Strict invariance** |  | Poor | 2793.8 (1066) ** | .037 | .047 | .94 | No |
| **2. SES**<br>**(all)** | 2039 (29) | Good | 4331.1 (71) | .026 | .050 | .99 |  |
| *High* | 802 (42) | Acceptable | 1056.5 (506) | .037 | .061 | .98 |  |
| *Middle* | 809 (37) | Acceptable | 1080.8 | .037 | .059 | .98 |  |
| *Low* | 423 (46) | Acceptable | 777.8 (506) | .036 | .068 | .98 |  |
| **1.Configural invariance** |  | Good | 3199.8 (1518) | .037 | .047 | .95 | Yes |
| **2.Metric invariance** |  | Good | 3199.8 (1518) | .037 | .047 | .95 | Yes |
| **3.Scalar invariance** |  | Good | 3310.1 (1572) * | .037 | .048 | .95 | No |
| **4.Strict invariance** |  | Good | 3383.5 (1626) * | .036 | .048 | .95 | No |

\* p = .05; \*\* p <.001. Models adjusted for grade, family type and immigration background

### Materials and procedure

Our research utilized the following variables from self-reported questionnaires:

#### Social Support Sources

The variables for social support sources were developed from fourteen self-reported items, which measured perceived support from family (four items), friends (four items), teachers (three items), and classmates (three items). The items were primarily sourced from the Multidimensional Scale of Perceived SS (MSPSS; Zimet et al., 1988) sub-scales, with a few specifically created for the HBSC survey. For instance:

Family Support: ‘My family really tries to help me’, ‘I get the emotional help and support I need from my family’, ‘I can talk to my family about my problems’, and ‘My family would like to help me make decisions’.

Friend Support: ‘My friends really try to help me’, ‘I can count on my friends when things go wrong’, ‘I have friends with whom I can share joy and sorrow’, ‘I can talk with my friends about my problems’.

Classmate Support: The items were drawn from the Teacher and Classmate Support Scale (Torsheim et al., 2012). Items were ‘Classmates enjoy being together’, ‘Classmates are kind/helpful’, and ‘Classmates accept me as I am’.

Teacher Support: These items were specifically crafted for the HBSC survey: ‘I feel that my teachers accept me as I am’, ‘I feel that my teachers care about me’, and ‘I feel that I can trust my teachers’.

Responses to classmate, teacher, and family SS items were marked on a 5-point Likert scale, ranging from 1 = ‘strongly disagree’ to 5 = ‘strongly agree’. Responses for SS from friends were graded on a 7-point Likert scale, where 1= ‘strongly disagree’ and 7 = ‘strongly agree’.

The scale’s structure was examined to discern whether it functioned as a single (Social Support) or multiple variables (Social Support from family, friends, teacher, and classmates). This was achieved via confirmatory factor analysis. The multifactorial structure was confirmed by comparing the multifactorial model fit (χ2=.000; CFI=.99; TLI= .99; RMSEA= .031; SRMR= .026; Alpha: Family= .88; Friends= .93; Classmates= .82; Teacher= 0.88; Omega: Family=.89; Friends= .93; Classmates= .82; Teacher= .88). In contrast, a unifactorial model did not achieve suitable model-based fit indexes (χ2=.000; CFI=.91; TLI= .89; RMSEA= .248; SRMR= .225; Cronbach’s alpha = .88; Omega = .90).

#### Mental Well-being

Mental well-being was evaluated using an adapted Danish version of the SWEMWBS (Stewart-Brown et al., 2009). The original brief version of WEMWBS, which was translated into Danish and validated in a Danish adult population (Koushede et al., 2019), was altered slightly by the Danish HBSC team for comprehensibility by children as young as 11. This adapted scale comprised seven items which asked participants how often they: ‘feel that things will go well in the future’, ‘feel useful’, ‘feel relaxed’, ‘solve problems well’, ‘think clearly’, ‘feel close to other people’, ‘have your own opinions’. In the HBSC questionnaire, responses were given on a 5-point scale, with scoring inverted so that higher scores indicated enhanced wellbeing (1 = never, 5 = always). This variable is managed in the analysis as a continuous variable with scores ranging from 7 to 35. The scale demonstrated good reliability in this sample with a Cronbach’s alpha of .83 and Omega of .84.

#### Problematic Social Media Use (PSMU)

Problematic social media use (PSMU) was assessed using the 9-item Social Media Disorder Scale (SMD scale) with a dichotomous (No/Yes) response format (Boer et al., 2021). The items, such as “I can not think of anything else”, “ I wish to spend more time”, “.I feel unwell when couldn’t use”, “I failed to spend less time”, “ I neglected other activities”, “I argued because of use”, “I lied about time spent”, “ I escape from negative feelings” and “I had conflict with family because of use” It. This was considered as a continuous variable, where a higher score implied higher problematic social media use. The scale’s reliability in this sample was satisfactory with a Cronbach’s alpha of .73 and Omega of .75.

#### Physical Activity

The physical activity level was assessed by three items:

1. Self-rated physical fitness: Participants rated their own physical fitness on a reversed 4-point scale, with 1=very good and 4=bad.
2. Frequency of vigorous physical activity outside school in the past week: Participants reported how often they had engaged in vigorous physical activity in the previous week, scored inversely from 1=every day to 7=never.
3. Duration of vigorous physical activity outside school in the past week: Participants reported the total time they spent on vigorous physical activity outside school in the previous week, with options ranging from 1=none to 5=about 4-6 hours per week.

The total score for the physical activity variable was calculated by summing the scores of these items (reversed when needed) and was treated as a continuous variable ranging from 3 to 16, with a higher score indicating a higher level of physical activity. The scale’s reliability in this sample was good with a Cronbach’s alpha of .82 and Omega of .85.

#### Socio-Economic Status (SES)

Family Ocupational Social Class (FOSC) is an appropriate indicator of the adolescents’ Socio-Economic Status (SES) (Pförtner et al. 2015) and it has been also used in previous studies with the same sample of Danish adolescents in the age group as this study (Holstein et al., 2020). Items composing this FOSC indicator were: ‘‘Does your father (mother) have a job?’’ ‘‘If yes, please write exactly what job he (she) does’’ ‘‘Please say in what place he (she) works.’’ The responses were coded according to the Danish Occupational Social Class measurement from I (high) to V (low). A category VI was added, including parents outside the labor market who receive unemployment benefits, disability pension or other kinds of transfer income (Christensen et al. 2014). Each participant was categorized by the highest-ranking parent into high (I–II), middle (III– IV) and low (V–VI) SES.

#### Covariates

The following sociodemographic variables were included as covariates in the analysis to control for their association with the sources of SS, mental well-being, PSMU and PA: (1) Grade (7 or 9), (2) immigration background: Danish origin (regardless of country of birth, having at least one parent born in Denmark); Descendant (born in Denmark to parents born outside of Denmark) and immigrant (born outside of Denmark to parents born outside of Denmark), (3) family type (traditional, single parent, reconstructed family, other type).

### Ethical considerations

The international HBSC research protocol specifies ethical principles for data collection and protection. In Denmark, there is no formal agency for approval of questionnaire-based surveys. To obtain approval and consent, the school board was asked as the parents’ representative, the headmaster, and the students’ council in each of the participating schools to approve the study. The participants received oral and written information that participation was voluntary, and that data were treated confidentially. The study complies with national standards for data protection. The Danish Data Protection Authority has requested notification of such studies and has granted acceptance for the 2018 survey (Case 10 622, University of Southern Denmark).

### Statistical analysis

Data analysis involved data visualization, descriptive analysis, and the parametric Mann-Whitney U test and the non-parametric Kruskal–Wallis’s test to examine differences in variable scores across gender and SES groups. To inspect the influences of different SS sources on adolescents’ mental well-being (MW), PSMU, and PA, and their respective associations, we used Structural Equation Modeling (SEM). Boxplots were utilized to evaluate data normality. Descriptive analyses (frequencies, means, Standard Deviations, Median, and Inter-quartile range) provided a summary of the data. The Mann-Whitney U test, a parametric test, was employed to identify differences in medians across gender groups for Family Support, Friends Support, Teacher Support, Classmate Support, Mental Well-being, PSMU, and Physical Activity variables. The non-parametric Kruskal–Wallis test was utilized to compare medians across SES groups.

SEM was employed to assess the influence of different sources of social support on the levels of adolescents’ mental well-being, problematic social media use, and physical activity, conceptualized as latent variables. These latent variables were composed of the items treated as observed variables. Confirmatory factor analysis (CFA) was carried out within SEM, and the measurement invariance of the model was subsequently tested to confirm group comparability. SEM provides a robust method for investigating complex dependencies within health and social sciences (Beran & Violato, 2010). This analysis allows for the investigation of latent variables and their relationships, thereby enabling the analysis of the dependencies of psychological constructs without measurement errors (Nachtigall et al., 2003). Furthermore, using a robust covariance-based comparison approach in multigroup analysis is a powerful way to investigate the effects of moderators on these associations (Raykov & Marcoulides, 2006).

During the modelling process, items were considered as ordinal variables to account for the non-normal distribution of the data. All missing data from the variables were deleted listwise, and ‘unclassified’ responses were treated as missing data. This was done using the Weighted Least Square Mean and Variance (WLSMV) estimator and pairwise-based imputation. The same analysis was carried out using different estimators to validate the consistency of the results. These included the Pairwise maximum likelihood (PML) estimator and Maximum likelihood with robust standard errors (MLR estimator) with variables treated as continuous, and Full Information Maximum likelihood (FIML).

For accurate group comparisons, configural, metric, and scalar invariances must be ensured, as measurement error can influence our ability to make accurate and meaningful comparisons (Vandenberg & Lance, 2000). Thus, testing measurement invariance provides a statistically rigorous way to ensure that comparisons represent true differences in our constructs of interest (Putnick & Bornstein, 2016). Measurement invariance was assessed by constraining the parameters step by step and using comparative fit indexes.

Initially, Configural invariance was aimed at examining the goodness of fit of the factor structure across the groups when factor loadings and item intercepts were unconstrained. This was done for both gender and SES groups. Next, the metric invariance was tested to examine if the factor loadings were equivalent across the groups by constraining each factor loading equally across groups. This aimed to inspect if each latent variable’s construct had the same meaning across groups. The third and fourth steps involved assessing scalar invariance to examine if the latent means can be compared across groups by themselves, with the item intercepts of the factors constrained as equal across groups in these steps (Milfont & Fisher, 2010). For each step of the confirmatory factor analysis (CFA), the chi-square statistic test was reported along with other fit indexes (CFI, RMSEA, SRMR) (see Table 3).

**Table 3.**
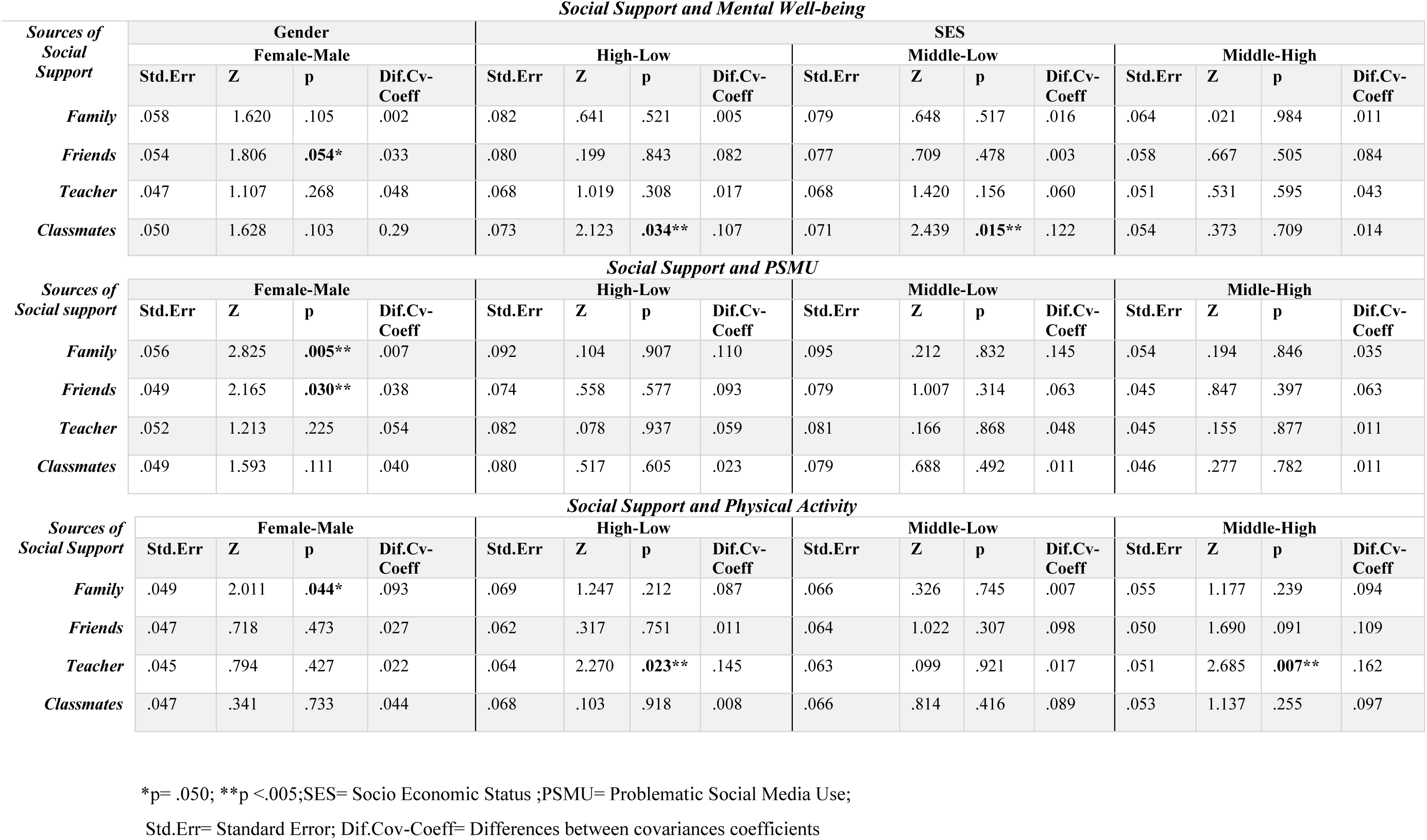
Covariance-based comparisons across Gender and SES groups.

Finally, upon calculating the structural model fit indexes and testing measurement invariance, a multigroup confirmatory factor analysis (MGCFA) was performed. Covariance-based comparisons were calculated to identify the potential moderation of gender and SES across the associations.

The software used was R (version 4.1.1) and the packages used were lavaan version 0.6-9 and semTools 0.5-5.

## 3. Results

### 3.1 Descriptive data and measurement invariance of the structural model

The present study utilized data from the 2018 Danish HBSC survey, incorporating a total of 2034 adolescents comprised of an even distribution of females (n = 1002; 49.3%) and males (n = 1032; 50.7%). Most participants (n = 1520; 74.7%) were part of a conventional family structure, consisting of a mother and father, while others belonged to single-parent households (n = 326; 16%), reconstructed families, i.e., living with a parent’s new partner (n = 154; 7.6%), and a small group (n = 27; 1.3%) didn’t identify with these categories.

In terms of immigration background, most of the sample (1825; 89.7%) had a Danish origin (at least one parent born in Denmark), regardless of the country of birth. A total of 6.4% (131) were children of immigrant parents (born in Denmark to parents born outside Denmark), 3.3% (68) were immigrants (born outside Denmark to parents also born outside Denmark), and 0.3% (6) didn’t fall into any of these categories. As for the Family Occupational Social Class (a marker of SES), the sample was skewed towards Middle-High SES (High= 802; 39.4%/ Middle= 809; 39.8%), with 423 (20%) belonging to a Low SES group.

Differences in total scores related to various sources of perceived Social Support, MWB, PSMU, and PA, across gender and SES were calculated. The results showed significant differences by gender and SES for all variables, excluding the scores for perceived friends support and MWB, which showed no statistically significant difference across gender or SES (refer to Table 1).

As outlined in the statistical analysis section, before making group comparisons, we conducted several tests of measurement invariance of the structural model across Gender and SES. The proposed structural equation model exhibited excellent goodness-of-fit indexes and an adequate measurement invariance across gender and SES groups, validating the structural model and covariances comparisons between groups (refer to Table 2). However, while the measurement invariance was confirmed at the configural and metric level, it was not achieved at the scalar level, the most restrictive comparison. This implies that comparing means or medians of these variables across gender and SES groups may not be statistically appropriate. Different groups might scale or range these variables differently, potentially leading to misinterpretation of results.

To facilitate comprehension, the following sections offer a qualitative summary of covariance-based comparisons between different social support sources and MWB, PSMU, and PA by gender and SES. However, all supporting quantitative information is available in Table 3. Information on associations between PSMU-MWB, PSMU-PA, and PA-MWB are also included in the text. Additionally, Figures 1 and 2 graphically represent the positive and negative associations between variables and their correlation coefficients.

**Figure 1.**
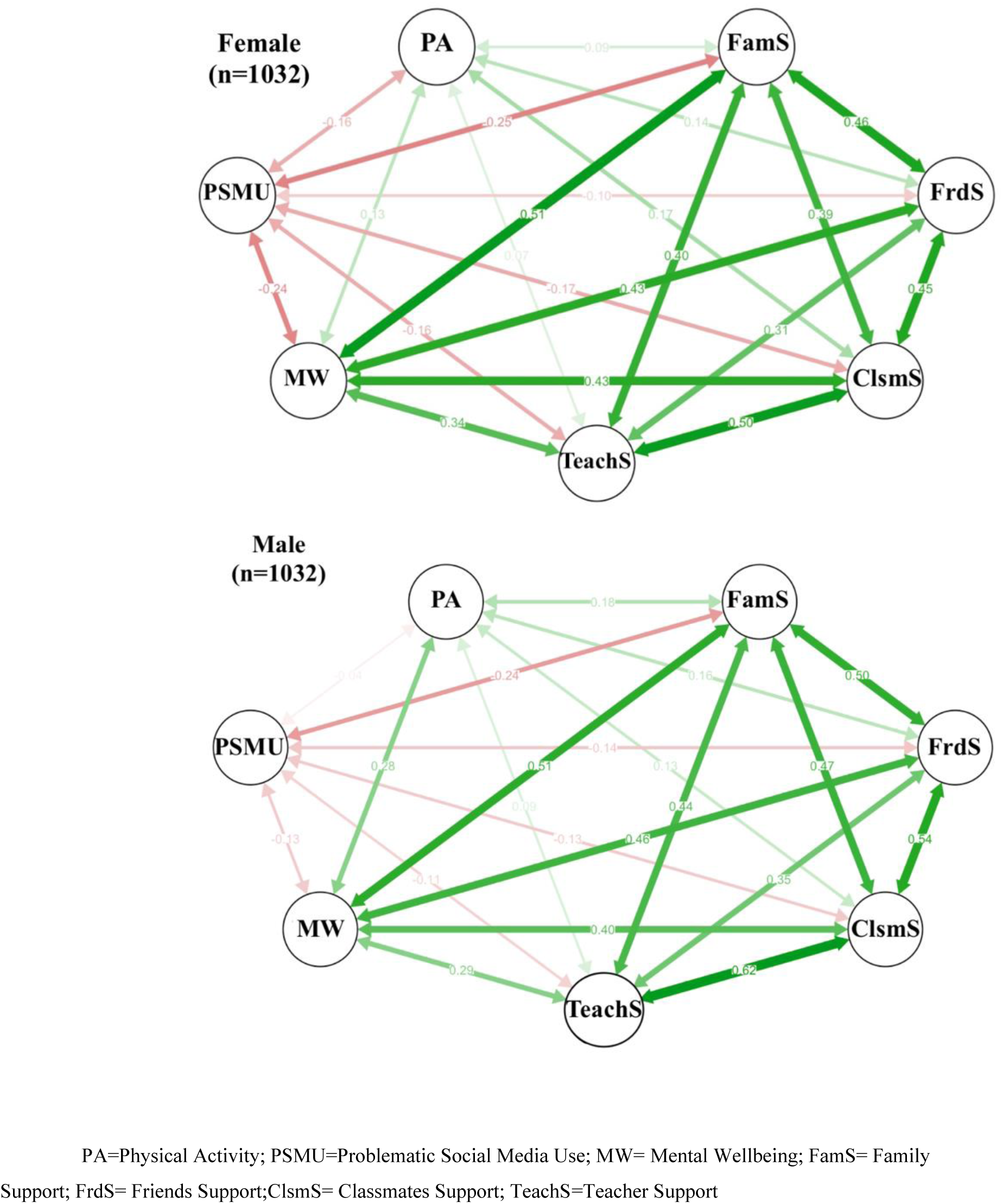
Gender based associations.

**Figure 2.**
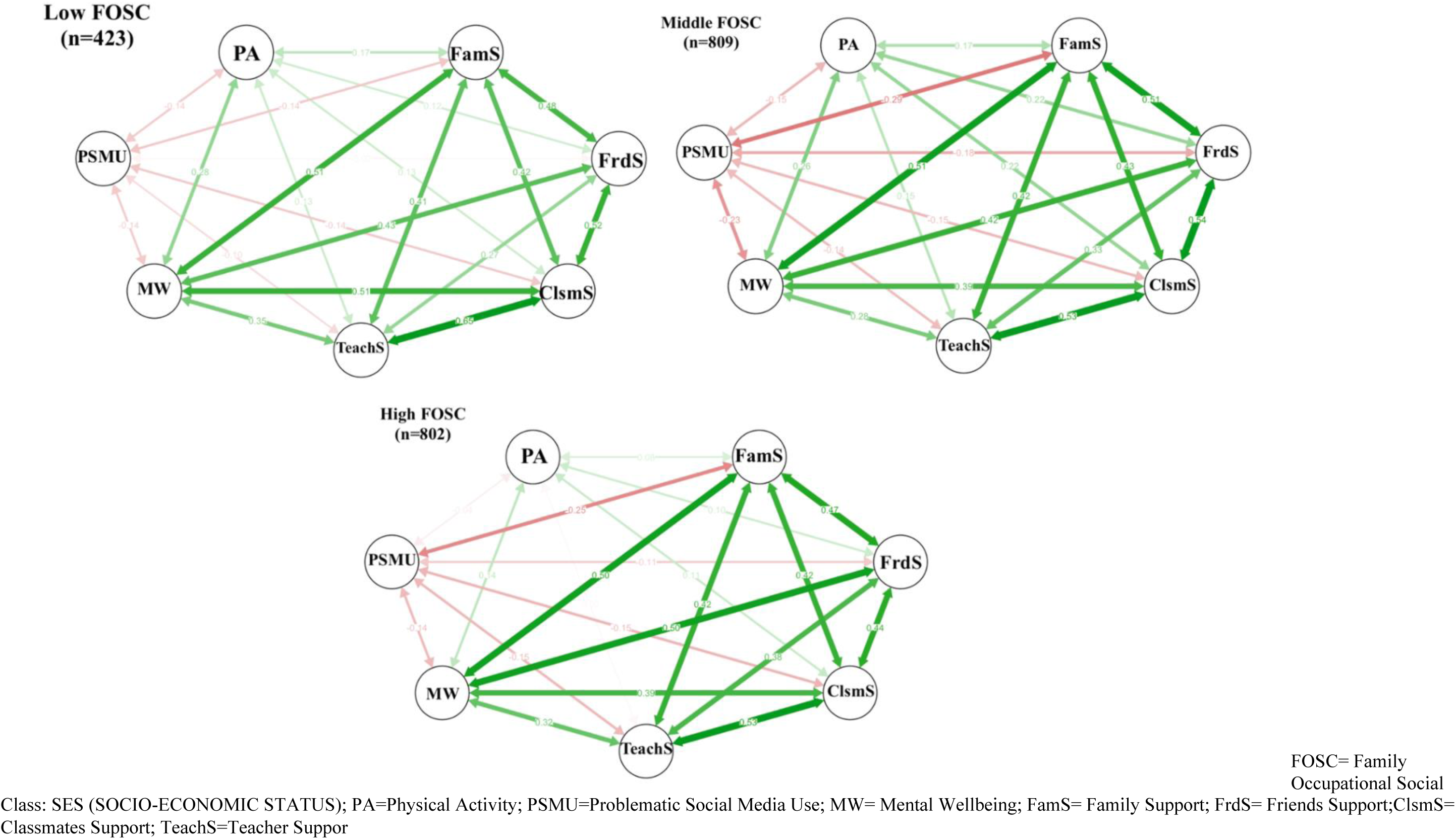
SES based associations.

### 3.2 Gender moderation

#### 3.2.1 Mental Wellbeing and Social Support sources

The positive association between MWB and various sources of perceived SS (Family, Friends, Teacher, and Classmates) showed no significant differences across genders.

#### 3.2.2 PSMU and Social Support sources

Significant differences were found for the negative association between PSMU and Family Support, which was stronger in females than males. In contrast, the negative association between PSMU and Friends Support was stronger in males than females. The negative associations between PSMU and both Teacher and Classmates support did not achieve statistical significance across genders.

#### 3.2.3 Physical Activity and Social Support sources

A more pronounced significant difference was found in the positive association between PA and Family Support, with the association being stronger in males than females. No significant differences were found regarding Friends, Teacher, or Classmates support.

#### 3.2.4 PSMU and Mental Well-being

We observed no significant gender-based differences in the correlation between PSMU and MWB (Std.Err= .059; Z= 1.068; p= .285; Dif.Cov-Coef= .115).

#### 3.2.5 PSMU and Physical Activity

We observed no significant gender-based differences in the correlation between PSMU and PA (Std.Err= .044; Z= .189; p=.850; Dif.Cov-Coef= .124).

#### 3.2.6 Physical Activity and MWB

A positive association was observed between MWB and PA, being significantly greater in males than females (Std.Err= .042; Z= 4.390; p<.001; Dif.Cov-Coef= .152).

### 3.3. Socio-Economic Status moderation

#### 3.3.1 Mental Wellbeing and Social Support sources

No statistically significant differences were found in the association between adolescents’ perceived family or friend support and mental wellbeing across SES groups. However, a stronger positive difference was found in the associations between classmates’ perceived support and adolescents’ Mental Well-being from low SES compared to those from Middle and High SES.

#### 3.3.2 PSMU and Social Support sources

No significant differences across SES groups were found regarding the negative association between different sources of social support and PSMU.

#### 3.3.3 Physical Activity and Social Support sources

For the positive associations between different sources of social support and PA, a statistically significant difference was found only between teacher support and PA, which was stronger in adolescents from Low-Middle SES compared to those from High SES.

#### 3.3.4 PSMU and Mental Well-being

No significant differences were found across SES for the relationship between PSMU and MWB (High vs Low: Std.err= .089; Z= .620; p=.535; Dif.Cov-Coeff= .014; Middle-vs Low : Std.err= .098; Z= .023; p=.981; Dif.Cov-Coeff= .095; Middle vs High: Std.err= .055; Z= 1.048; p=.294; Dif.Cov-Coeff= .081).

#### 3.3.5 PSMU and Physical Activity

No SES-based moderation was found for the association between PSMU and PA (High vs Low: Std.err= .066; Z= 1.620; p=.105; Dif.Cov-Coeff= .088; Middle-vs Low : Std.err= .071; Z= .597; p=.551; Dif.Cov-Coeff= .024; Middle vs High: Std.err= .048; Z= 1.340; p=.180; Dif.Cov-Coeff= .112).

#### 3.3.6 Physical Activity and Mental Well-being

In terms of differences across SES for the association between MWB and PA, a stronger positive relationship was found in groups from low SES (Std.err= .063; Z= 2.295; p=.022; Dif.Cov-Coeff= .131) and middle SES (Std.err= .052; Z=2.127; p=.033; Dif.Cov-Coef= .126) compared to those from the High SES.

## 4. Discussion

This study aimed to investigate the potential moderating effects of gender and socio-economic status (SES) on the relationships among various sources of social support, mental well-being (MWB), problematic social media use (PSMU), and physical activity (PA). The findings provide substantial support for the positive relationship between social support and both MWB and PA. Additionally, negative relationships were found between social support and PSMU. Confirming earlier hypotheses, the study uncovered negative associations between PSMU and MWB and PA and positive associations between PA and MWB.

However, the moderating role of gender was only confirmed in specific associations, namely, PSMU-Family and PSMU-Friend Support. Meanwhile, socio-economic status emerged as a significant moderator only in MWB-Teacher Support, MWB-Classmate Support, and PA-Teacher Support. The interaction between PA and MWB was found to be moderated by both gender and SES.

It is noteworthy that the rest of the examined associations did not exhibit significant disparities across gender or SES. This paper explores these results further, aiming to shed light on how gender and SES may influence these complex interrelationships.

Interestingly, the strong positive link between MWB and support from teachers or classmates in adolescents from low SES backgrounds mirrors previous studies emphasizing the pivotal role school-based social support plays in promoting adolescents’ mental well-being (Delaruelle et al., 2021; McPherson et al., 2014). It appears that schools can serve as critical environments for fostering social capital, which in turn, contributes significantly to the mental health of children and young people (Nielsen et al., 2015).

Regarding the negative association between PSMU and social support sources, our results highlight that the role of gender was more pronounced than that of SES. The findings suggest that familial support may be particularly influential in mitigating PSMU in girls, while boys might derive more significant benefits from friends’ support. This resonates with earlier studies, such as Marengo et al. (2021), showing that girls often report higher PSMU than boys, but also that social support can attenuate the risk of PSMU. These studies, however, did not differentiate between support sources. Consequently, the differences in gender moderation we observed might be attributed to girls receiving less friend support but more familial support than boys.

In contrast, previous studies (e.g., Borrachino et al., 2022; Del Rey et al., 2019) conducted with Italian and Spanish adolescents have suggested that social support from school, including teachers and classmates, can provide significant protection against PSMU, irrespective of gender differences. This suggests that the protective influence of specific social support sources on PSMU may vary across different samples and countries. This calls for more research exploring these differences.

Our findings can be contextualized in the broader literature. For instance, Jakobsen et al. (2022) particularly beneficial for adolescents from low and mid SES backgrounds to engage in physical activities compared to those from high SES backgrounds.

Earlier studies have often reported boys engaging in higher levels of PA than girls (Telford et al., 2016; Hu et al., 2021). Our results hint that this may be driven, at least in part, by differing perceptions of familial support among boys and girls. As Reimers et al. (2019) noted, while social support generally correlates with higher participation in physical activities, it might be essential to boost familial support for girls to reap the same benefits that boys do.

When considering SES differences, the role of school appears to be crucial in the association between social support and physical activity, regardless of gender. As our study showed, teacher support might be especially impactful for adolescents from low SES backgrounds. This is in line with studies such as Rajbhandari-Thapa et al. (2022) and Alliott et al. (2022), which highlighted the positive influence of supportive school environments on PA among adolescents from low-SES backgrounds.

In regards to the positive relationship between PA and MWB, it was both moderated by gender and SES, showing that boys could benefit more from PA in terms of MWB than girls. And also, that adolescents from low -mid SES may get greater benefit on their MWB from PA participation when compared to adolescents from highs SES. As previous evidence has showed, adolescents from high SES were more likely to participate in sport, both in frequency and duration (Owen et al., 2022) and that being physically active lifestyles during childhood and adolescence is associated with better mental health (Bull et al., 2020), perhaps due to a higher family support as out data are suggesting.

Similarly, previous literature has discussed associations from Social Support and PA across SES groups, where these socio-economic disparities could be due to different facilitators and barriers experienced across adolescents with different socio-economic backgrounds (Alliot et al, 2022). According to Bull et al. (2022) children and adolescents from families with high-SES might experience parental encouragement or pressure to priorities academic tasks, rather than leisure activities. Meanwhile, adolescents from families with low SES may face additional barriers to structured sports, such as the associated financial costs (e.g., registration fees, equipment), transportation issues, and limited or poor availability of quality facilities and activities in the local neighborhood and at school (Alliot et al., 2022).Therefore, while adolescents from high and low SES experience different barriers across different domains of PA participation, contributing to differential benefits from this physical activity participation on their mental well-being, school support can be critical to ensure PA participation in all groups.

Lastly, the negative association between PSMU-MWB, as well as PSMU-PA were not moderated by non-significant associations were detected across gender and SES groups, which is indicating that the detrimental effect of PSMU on adolescents’ MWB and PA are the same across gender and SES groups. However, this negative association should be considered in future studies covering mediation role of PSMU in the relationship between SS sources and MWB. For example, those who experience a poor Social Support might report low levels of mental well-being and consequently, may become vulnerable social media users as a coping strategy to regulate their emotional and social difficulties putting themselves at major risk to develop a maladaptive use of social media and therefore, decreasing their mental well-being consequently (Satici & Uysal, 2015; Hong, Huang, Lin & Chiu, 2014). But also, as previous studies have noted, PSMU could be also considered as an independent variable, where a mood modification, cognitive preoccupation, and compulsive online behaviors, might result in negative consequences in terms of emotional, social, and school impairments and therefore being the predictor of less social support perception (Keles, MacCraen & Gralish, 2020; Marion et al., 2020).

### Strengths and limitations

A main strength of this study is the large, current, and nationally representative dataset from the HBSC international project, which entails that the results can be generalized to the wider Danish adolescent population. Another important strength is the beneficial study of investigating the association between perceived SS from various sources, such as friends, parents and teachers, and several indicators of mental health, considering the measurement errors of latent constructs rather than observed variables. In addition, our study included several important possible confounders, such as grade, family type and immigration background that may influence these relationships.

Nevertheless, some limitations need to be highlighted as the analyses were necessarily cross-sectional, meaning that causal relations cannot be inferred from these results alone and data collection was based on self-reports only from a unique source of information.

## 5. Conclusions and future implications

This study offers the opportunity to understand better simultaneous associations between indicators of adolescents’ behavioral and mental health indicators (SS, MWB, PSMU and PA) that have received major attention nowadays either by clinicals and academics and show the comparative effect of social determinants as gender and SES in these relations.

These results are of major importance as a first step for future studies aimed to model more complex mediated relationships between social support sources and MWB, PSMU and PA. For example, further research is required to investigate those relations longitudinally in order understand whether different sources of perceived SS would predict later MWB, PSMU or PA. It would also be relevant to examine the other way around with MWB, exploring how MWB jointly with other socio-demographic factors would predict different sources of perceived SS, levels of PSMU or PA. Whereas, exploring the mediating role of PSMU and PA in the relationship between SS sources and MWB would be also of great interest.

Similarly, future studies should be conducted in different populations and specific groups, to test whether same associations and differences can be identified and compared between different groups, not only based on socio-demographic characteristics but also, on youth with different profiles of MWB, PSM users or PA and so, to be able to identify who could benefit the most for targeted interventions.

In terms of practical implications, findings support the importance to ensure different sources of social support to promote mental health in adolescents and prevent health behaviors problems as problematic use of technologies and the detrimental consequences of sedentarism. Meanwhile, at the targeted levels, effective interventions aimed to provide family support can be critical to prevent PSMU especially in girls, whereas school social support resources can be more beneficial to improve mental wellbeing and PA in adolescents from more economically disadvantaged families. Therefore, public health policies and intervention fostering family and school support can help protect adolescents’ mental health, reducing the risk of problematic media use and lack of physical activity participation.

## Author Contributions

Three researchers (E.B.M., Z.I.S. and KRM.) participated in the first conceptualization of the study and first drafting of the manuscript. Three researchers participated in methodology: data analysis and interpretation of data (E.B.M., M.T.S and Z.I.S.). Four researchers participated in writing—review and editing (E.B.M, K.R.M., Z.I.S and M.T.S). All eight authors have contributed to revision of the paper for intellectual content and final approval of the version: E.B.M; K.R.M, M.T.S, Z.I.S, M.T.D, L.N, C.B.M, M.T.

Data and HTML file with procedures of data analysis and codes are available upon request.

## Data Availability

All data produced in the present study are available upon reasonable request to the authors. HTML file with all the analysis procedures and codes can be requested

## References

1. Alliott, O., Ryan, M., Fairbrother, H., & Van Sluijs, E. (2022). Do adolescents’ experiences of the barriers to and facilitators of physical activity differ by socioeconomic position? A systematic review of qualitative evidence. Obesity Reviews, 23(3), e13374.

2. Bell, S. L., Audrey, S., Gunnell, D., Cooper, A., & Campbell, R. (2019). The relationship between physical activity, mental wellbeing and symptoms of mental health disorder in adolescents: a cohort study. International Journal of Behavioral Nutrition and Physical Activity, 16(1), 1–12.

3. Beran, T. N., & Violato, C. (2010). Structural equation modeling in medical research: a primer. BMC Research Notes, 3(1), 1–10.

4. Biddle, S. J. H., & Asare, M. (2011). Physical activity and mental health in children and adolescents: a review of reviews. British Journal of Sports Medicine, 45(11), 886–895.

5. Boer, M., Stevens, G. W. J. M., Finkenauer, C., Koning, I. M., & van den Eijnden, R. J. J. M. (2021). Validation of the social media disorder scale in adolescents: Findings from a large-scale nationally representative sample. Assessment, 10731911211027232.

6. Boer, M., Van Den Eijnden, R. J. J. M., Boniel-Nissim, M., Wong, S.-L., Inchley, J. C., Badura, P., Craig, W. M., Gobina, I., Kleszczewska, D., & Klanšček, H. J. (2020). Adolescents’ intense and problematic social media use and their well-being in 29 countries. Journal of Adolescent Health, 66(6), S89–S99.

7. Boer, M., van den Eijnden, R. J. J. M., Finkenauer, C., Boniel-Nissim, M., Marino, C., Inchley, J., Cosma, A., Paakkari, L., & Stevens, G. W. J. M. (2022). Cross-national validation of the social media disorder scale: findings from adolescents from 44 countries. Addiction, 117(3), 784–795.

8. Boniel-Nissim, M., van den Eijnden, R. J. J. M., Furstova, J., Marino, C., Lahti, H., Inchley, J., Šmigelskas, K., Vieno, A., & Badura, P. (2022). International perspectives on social media use among adolescents: Implications for mental and social well-being and substance use. Computers in Human Behavior, 129, 107144.

9. Bor, W., Dean, A. J., Najman, J., & Hayatbakhsh, R. (2014). Are child and adolescent mental health problems increasing in the 21st century? A systematic review. Australian & New Zealand Journal of Psychiatry, 48(7), 606–616.

10. Borraccino, A., Marengo, N., Dalmasso, P., Marino, C., Ciardullo, S., Nardone, P., … & 2018 HBSC-Italia Group. (2022). Problematic social media use and cyber aggression in Italian adolescents: the remarkable role of social support. International journal of environmental research and public health, 19(15), 9763.

11. Brooks, F., Zaborskis, A., Tabak, I., Carmen Granado Alcón, M. del, Zemaitiene, N., de Roos, S., & Klemera, E. (2015). Trends in adolescents’ perceived parental communication across 32 countries in Europe and North America from 2002 to 2010. The European Journal of Public Health, 25(suppl_2), 46–50.

12. Buda, G., Lukoševičiūtė, J., Šalčiūnaitė, L., & Šmigelskas, K. (2021). Possible effects of social media use on adolescent health behaviors and perceptions. Psychological Reports, 124(3), 1031–1048.

13. Bull FC, Al-Ansari SS, Biddle S, Borodulin K, Buman MP, Cardon G, et al. World Health Organization 2020 guidelines on physical activity and sedentary behaviour. Br J Sports Med. 2020;54(24):1451–62.

14. Cheng, S.-T., & Chan, A. C. M. (2004). The multidimensional scale of perceived social support: dimensionality and age and gender differences in adolescents. Personality and Individual Differences, 37(7), 1359–1369.

15. Christensen, U., Krølner, R., Nilsson, C. J., Lyngbye, P. W., Hougaard, C. Ø., Nygaard, E., Thielen, K., Holstein, B. E., Avlund, K., & Lund, R. (2014). Addressing social inequality in aging by the Danish occupational social class measurement. Journal of Aging and Health, 26(1), 106–127.

16. Chu, P. Sen, Saucier, D. A., & Hafner, E. (2010). Meta-analysis of the relationships between social support and well-being in children and adolescents. Journal of Social and Clinical Psychology, 29(6), 624.

17. Delaruelle, K., Walsh, S. D., Dierckens, M., Deforche, B., Kern, M. R., Currie, C., … & Stevens, G. W. (2021). Mental health in adolescents with a migration background in 29 European countries: the buffering role of social capital. Journal of Youth and Adolescence, 50(5), 855–871.

18. Del Rey, R. Ortega-Ruiz, J.A. Casas Asegúrate: an intervention program against cyberbullying based on teachers’ commitment and on design of its instructional materialsInt J Environ Res Publ Health, 16 (3)

19. Franchina, V., Vanden Abeele, M., Van Rooij, A. J., Lo Coco, G., & De Marez, L. (2018). Fear of missing out as a predictor of problematic social media use and phubbing behavior among Flemish adolescents. International Journal of Environmental Research and Public Health, 15(10), 2319.

20. Fusar-Poli P, Correll CU, Arango C, Berk M, Patel V, Ioannidis JPA. Preventive psychiatry: a blueprint for improving the mental health of young people. World Psychiatry. 2021 Jun;20(2):200–221. doi: 10.1002/wps.20869. PMID: 34002494; PMCID: PMC8129854.

21. Gariepy, G., Honkaniemi, H., & Quesnel-Vallee, A. (2016). Social support and protection from depression: systematic review of current findings in Western countries. The British Journal of Psychiatry, 209(4), 284–293.

22. Gomes, A. C., Rebelo, M. A. B., de Queiroz, A. C., de Queiroz Herkrath, A. P. C., Herkrath, F. J., Rebelo Vieira, J. M., … & Vettore, M. V. (2020). Socioeconomic status, social support, oral health beliefs, psychosocial factors, health behaviours and health-related quality of life in adolescents. Quality of life research, 29(1), 141–151.

23. Heerde, J. A., & Hemphill, S. A. (2018). Examination of associations between informal help-seeking behavior, social support, and adolescent psychosocial outcomes: A meta-analysis. Developmental Review, 47, 44–62.

24. Holstein, B. E., Damsgaard, M. T., Madsen, K. R., & Rasmussen, M. (2020). Persistent social inequality in low life satisfaction among adolescents in Denmark 2002–2018. Children and Youth Services Review, 116, 105097.

25. Hong, F.-Y., Huang, D.-H., Lin, H.-Y., & Chiu, S.-L. (2014). Analysis of the psychological traits, Facebook usage, and Facebook addiction model of Taiwanese university students. Telematics and Informatics, 31(4), 597–606.

26. Hu, D., Zhou, S., Crowley-McHattan, Z. J., & Liu, Z. (2021). Factors that influence participation in physical activity in school-aged children and adolescents: a systematic review from the social ecological model perspective. International Journal of Environmental Research and Public Health, 18(6), 3147.

27. Humbert, M. L., Chad, K. E., Spink, K. S., Muhajarine, N., Anderson, K. D., Bruner, M. W., Girolami, T. M., Odnokon, P., & Gryba, C. R. (2006). Factors that influence physical activity participation among high-and low-SES youth. Qualitative Health Research, 16(4), 467–483.

28. Inchley, J., Currie, D., Budisavljevic, S., Torsheim, T., Jåstad, Cosma, A., Kelly, C., & Arnarsson, A.M. (Eds.) (2020) Spotlight on adolescent health and well-being: Findings from the 2017/18 Health Behaviour in School-aged Children (HBSC) Survey in Europe and Canada. https://apps.who.int/iris/bitstream/handle/10665/332091/9789289055000-eng.pdf

29. Ivie, E. J., Pettitt, A., Moses, L. J., & Allen, N. B. (2020). A meta-analysis of the association between adolescent social media use and depressive symptoms. Journal of Affective Disorders, 275, 165–174.

30. Jakobsen, A. L., Hansen, C. D., & Andersen, J. H. (2022). The association between perceived social support in adolescence and positive mental health outcomes in early adulthood: a prospective cohort study. Scandinavian Journal of Public Health, 50(3), 404–411.

31. Jusienė, R., Breidokienė, R., Sabaliauskas, S., Mieziene, B., & Emeljanovas, A. (2022). The Predictors of Psychological Well-Being in Lithuanian Adolescents after the Second Prolonged Lockdown Due to COVID-19 Pandemic. International Journal of Environmental Research and Public Health, 19(6), 3360.

32. Keles, B., McCrae, N., & Grealish, A. (2020). A systematic review: the influence of social media on depression, anxiety and psychological distress in adolescents. International Journal of Adolescence and Youth, 25(1), 79–93.

33. Kelly, Y., Zilanawala, A., Booker, C., & Sacker, A. (2018). Social media use and adolescent mental health: Findings from the UK Millennium Cohort Study. EClinicalMedicine, 6, 59– 68.

34. Khan, A., Lee, E.-Y., Janssen, I., & Tremblay, M. S. (2022). Associations of Passive and Active Screen Time With Psychosomatic Complaints of Adolescents. American Journal of Preventive Medicine.

35. Kircaburun, K., Griffiths, M. D., & Billieux, J. (2020). Childhood emotional maltreatment and problematic social media use among adolescents: The mediating role of body image dissatisfaction. International Journal of Mental Health and Addiction, 18(6), 1536–1547.

36. Koushede, V., Lasgaard, M., Hinrichsen, C., Meilstrup, C., Nielsen, L., Rayce, S. B., Torres-Sahli, M., Gudmundsdottir, D. G., Stewart-Brown, S., & Santini, Z. I. (2019). Measuring mental well-being in Denmark: Validation of the original and short version of the Warwick-Edinburgh mental well-being scale (WEMWBS and SWEMWBS) and cross-cultural comparison across four European settings. Psychiatry Research, 271, 502–509.

37. Koushede, V., & Donovan, R. (2022). Applying salutogenesis in community-wide mental health promotion. In The handbook of salutogenesis (pp. 479–490). Springer, Cham.

38. Kross, E., Verduyn, P., Sheppes, G., Costello, C. K., Jonides, J., & Ybarra, O. (2021). Social media and well-being: Pitfalls, progress, and next steps. Trends in Cognitive Sciences, 25(1), 55–66.

39. Leather, N. C. (2009). Risk-taking behaviour in adolescence: a literature review. Journal of Child Health Care, 13(3), 295–304.

40. McPherson, K. E., Kerr, S., McGee, E., Morgan, A., Cheater, F. M., McLean, J., & Egan, J. (2014). The association between social capital and mental health and behavioural problems in children and adolescents: an integrative systematic review. BMC psychology, 2(1), 1–16.

41. Maheswari, K., & Martin, M. R. M. (2022). Assessment of Socio-Economic Status and Mental Health of Adolescent School Girls. Journal of Positive School Psychology, 2805–2812.

42. Mahon, N. E., & Yarcheski, A. (2017). Parent and friend social support and adolescent hope. Clinical Nursing Research, 26(2), 224–240.

43. Maenhout, L., Peuters, C., Cardon, G., Compernolle, S., Crombez, G., & DeSmet, A. (2020). The association of healthy lifestyle behaviors with mental health indicators among adolescents of different family affluence in Belgium. BMC Public Health, 20(1), 1–13.

44. Marino, C., Gini, G., Angelini, F., Vieno, A., & Spada, M. M. (2020). Social norms and e-motions in problematic social media use among adolescents. Addictive Behaviors Reports, 11, 100250.

45. Marino, C., Gini, G., Vieno, A., & Spada, M. M. (2018). The associations between problematic Facebook use, psychological distress and well-being among adolescents and young adults: A systematic review and meta-analysis. Journal of Affective Disorders, 226, 274–281.

46. Marino, C., Lenzi, M., Canale, N., Pierannunzio, D., Dalmasso, P., Borraccino, A., Cappello, N., Lemma, P., & Vieno, A. (2020). Problematic social media use: associations with health complaints among adolescents. Annali Dell’Istituto Superiore Di Sanità, 56(4), 514–521.

47. Marengo, N., Borraccino, A., Charrier, L., Berchialla, P., Dalmasso, P., Caputo, M., & Lemma, P. (2021). Cyberbullying and problematic social media use: an insight into the positive role of social support in adolescents—data from the Health Behaviour in School-aged Children study in Italy. Public Health, 199, 46–50.

48. Mendonça, G., Cheng, L. A., Mélo, E. N., & de Farias Junior, J. C. (2014). Physical activity and social support in adolescents: a systematic review. Health Education Research, 29(5), 822– 839.

49. Milfont, T. L., & Fischer, R. (2010). Testing measurement invariance across groups: Applications in cross-cultural research. International Journal of Psychological Research, 3(1), 111–130.

50. Micheli, M. (2016). Social networking sites and low-income teenagers: between opportunity and inequality. Information, Communication & Society, 19(5), 565–581.

51. Nachtigall, C., Kroehne, U., Funke, F., & Steyer, R. (2003). Pros and cons of structural equation modeling. Methods Psychological Research Online, 8(2), 1–22.

52. Nagy-Pénzes, G., Vincze, F., & Bíró, É. (2020). Contributing factors in adolescents’ mental well-being:the role of socioeconomic status, social support, and health behavior. Sustainability, 12(22), 9597.

53. Nielsen, L., Koushede, V., Vinther-Larsen, M., Bendtsen, P., Ersbøll, A. K., Due, P., & Holstein, B. E. (2015). Does school social capital modify socioeconomic inequality in mental health? A multi-level analysis in Danish schools. Social Science & Medicine, 140, 35–43.

54. Oberst, U., Wegmann, E., Stodt, B., Brand, M., & Chamarro, A. (2017). Negative consequences from heavy social networking in adolescents: The mediating role of fear of missing out. Journal of Adolescence, 55, 51–60.

55. Odgers, C. L., & Jensen, M. R. (2020). Annual Research Review: Adolescent mental health in the digital age: facts, fears, and future directions. Journal of Child Psychology and Psychiatry, 61(3), 336–348.

56. Oliveira, J. M. D. de, Butini, L., Pauletto, P., Lehmkuhl, K. M., Stefani, C. M., Bolan, M., Guerra, E., Dick, B., De Luca Canto, G., & Massignan, C. (2022). Mental health effects prevalence in children and adolescents during the COVID-19 pandemic: A systematic review. Worldviews on Evidence-Based Nursing, 19(2), 130–137.

57. Owen, K. B., Nau, T., Reece, L. J., Bellew, W., Rose, C., Bauman, A., … & Smith, B. J. (2022). Fair play? Participation equity in organised sport and physical activity among children and adolescents in high income countries: a systematic review and meta-analysis. International Journal of Behavioral Nutrition and Physical Activity, 19(1), 1–13.

58. Paakkari, L., Tynjälä, J., Lahti, H., Ojala, K., & Lyyra, N. (2021). Problematic social media use and health among adolescents. International Journal of Environmental Research and Public Health, 18(4), 1885.

59. Pförtner, T.-K., Günther, S., Levin, K. A., Torsheim, T., & Richter, M. (2015). The use of parental occupation in adolescent health surveys. An application of ISCO-based measures of occupational status. J Epidemiol Community Health, 69(2), 177–184.

60. Potrebny, T., Wiium, N., & Lundegård, M. M.-I. (2017). Temporal trends in adolescents’ self-reported psychosomatic health complaints from 1980-2016: A systematic review and meta-analysis. PLOS One, 12(11), e0188374.

61. Putnick, D. L., & Bornstein, M. H. (2016). Measurement invariance conventions and reporting: The state of the art and future directions for psychological research. Developmental Review, 41, 71–90.

62. Khan, A., Lee, E. Y., Rosenbaum, S., Khan, S. R., & Tremblay, M. S. (2021). Dose-dependent and joint associations between screen time, physical activity, and mental wellbeing in adolescents: an international observational study. The Lancet Child & Adolescent Health, 5(10), 729–738.

63. Rajbhandari-Thapa, J., Metzger, I., Ingels, J., Thapa, K., & Chiang, K. (2022). School climate-related determinants of physical activity among high school girls and boys. Journal of Adolescence.

64. Raudsepp, L., & Kais, K. (2019). Longitudinal associations between problematic social media use and depressive symptoms in adolescent girls. Preventive Medicine Reports, 15, 100925.

65. Raykov, T., & Marcoulides, G. A. (2012). A first course in structural equation modeling. routledge.

66. Reimers, A. K., Schmidt, S. C., Demetriou, Y., Marzi, I., & Woll, A. (2019). Parental and peer support and modelling in relation to domain-specific physical activity participation in boys and girls from Germany. PLoS One, 14(10), e0223928.

67. Twenge, J. M., Joiner, T. E., Rogers, M. L., & Martin, G. N. (2018). Increases in Depressive Symptoms, Suicide-Related Outcomes, and Suicide Rates Among U.S. Adolescents After 2010 and Links to Increased New Media Screen Time. Clinical Psychological Science, 6(1), 3–17. 10.1177/2167702617723376

68. Samji, H., Wu, J., Ladak, A., Vossen, C., Stewart, E., Dove, N., Long, D., & Snell, G. (2022). Mental health impacts of the COVID-19 pandemic on children and youth–a systematic review. Child and Adolescent Mental Health, 27(2), 173–189.

69. Sanz-Martín, D., Melguizo-Ibáñez, E., Ruiz-Tendero, G., & Ubago-Jiménez, J. L. (2022). An Explanatory Model of the Relationships between Physical Activity, Social Support and Screen Time among Adolescents. International Journal of Environmental Research and Public Health, 19(12), 7463.

70. Satici, S. A., & Uysal, R. (2015). Well-being and problematic Facebook use. Computers in Human Behavior, 49, 185–190.

71. Scott, H., & Woods, H. C. (2018). Fear of missing out and sleep: Cognitive behavioural factors in adolescents’ nighttime social media use. Journal of Adolescence, 68, 61–65.

72. Stalsberg R, Pedersen AV. Effects of socioeconomic status on the physical activity in adolescents: a systematic review of the evidence. Scand J Med Sci Sports. 2010;20(3):368–83.

73. Telama, R., Laakso, L., Nupponen, H., Rimpelä, A., & Pere, L. (2009). Secular trends in youth physical activity and parents’ socioeconomic status from 1977 to 2005. Pediatric Exercise Science, 21(4).

74. Telford, R. M., Telford, R. D., Olive, L. S., Cochrane, T., & Davey, R. (2016). Why are girls less physically active than boys? Findings from the LOOK Longitudinal study. PLoS ONE, 11(3), e0150041. 10.1371/journal.pone.0150041

75. Torsheim, T., Samdal, O., Rasmussen, M., Freeman, J., Griebler, R., & Dür, W. (2012). Cross-national measurement invariance of the teacher and classmate support scale. Social Indicators Research, 105(1), 145–160.

76. Van Den Eijnden, R., Koning, I., Doornwaard, S., Van Gurp, F., & Ter Bogt, T. (2018). The impact of heavy and disordered use of games and social media on adolescents’ psychological, social, and school functioning. Journal of Behavioral Addictions, 7(3), 697– 706.

77. Van den Eijnden, R. J., Geurts, S. M., Ter Bogt, T. F., van der Rijst, V. G., & Koning, I. M. (2021). Social media use and adolescents’ sleep: A longitudinal study on the protective role of parental rules regarding internet use before sleep. International journal of environmental research and public health, 18(3), 1346.

78. Van Droogenbroeck, F., Spruyt, B., & Keppens, G. (2018). Gender differences in mental health problems among adolescents and the role of social support: results from the Belgian health interview surveys 2008 and 2013. BMC psychiatry, 18(1), 1–9.

79. Vandenberg, R. J., & Lance, C. E. (2000). A review and synthesis of the measurement invariance literature: Suggestions, practices, and recommendations for organizational research. Organizational Research Methods, 3(1), 4–70.

80. Veselska, Z., Madarasova Geckova, A., Gajdosova, B., Orosova, O., van Dijk, J. P., & Reijneveld, S. A. (2010). Socio-economic differences in self-esteem of adolescents influenced by personality, mental health and social support. European Journal of Public Health, 20(6), 647–652.

81. Walsh, S. D., Sela, T., De Looze, M., Craig, W., Cosma, A., Harel-Fisch, Y., Boniel-Nissim, M., Malinowska-Cieślik, M., Vieno, A., & Molcho, M. (2020). Clusters of contemporary risk and their relationship to mental well-being among 15-year-old adolescents across 37 countries. Journal of Adolescent Health, 66(6), S40–S49.

82. Wartberg, L., Kriston, L., & Thomasius, R. (2020). Internet gaming disorder and problematic social media use in a representative sample of German adolescents: Prevalence estimates, comorbid depressive symptoms and related psychosocial aspects. Computers in Human Behavior, 103, 31–36.

83. Woodfield, L., Duncan, M., Al-Nakeeb, Y., Nevill, A., & Jenkins, C. (2002). Sex, ethnic and socio-economic differences in children’s physical activity. Pediatric Exercise Science, 14(3), 277–285.

84. Worsley, J. D., McIntyre, J. C., Bentall, R. P., & Corcoran, R. (2018). Childhood maltreatment and problematic social media use: The role of attachment and depression. Psychiatry Research, 267, 88–93.

85. Yoon, Y., Eisenstadt, M., Lereya, S. T., & Deighton, J. (2022). Gender difference in the change of adolescents’ mental health and subjective wellbeing trajectories. European Child & Adolescent Psychiatry, 1–10.

86. Zhang, J., Marino, C., Canale, N., Charrier, L., Lazzeri, G., Nardone, P., & Vieno, A. (2022). The Effect of Problematic Social Media Use on Happiness among Adolescents: The Mediating Role of Lifestyle Habits. International Journal of Environmental Research and Public Health, 19(5), 2576.

87. Zimet, G. D., Dahlem, N. W., Zimet, S. G., & Farley, G. K. (1988). The multidimensional scale of perceived social support. Journal of Personality Assessment, 52(1), 30–41.

